# Is it inside my head? Characterization of sound externalization in schizophrenia

**DOI:** 10.1101/2025.10.08.25337491

**Authors:** Laure Fivel, Mathieu Lavandier, Nicolas Grimault, Fabien Perrin, Marine Mondino, Frédéric Haesebaert

## Abstract

Schizophrenia has been linked to reality monitoring confusions, particularly misattributions of internal productions to external sources. We hypothesized that these misattributions may be related to deficits in processing acoustic cues that distinguish the subjective experience of a sound source inside or outside the head, i.e., sound externalization. This study aimed to investigate sound externalization in patients with schizophrenia, particularly for emotional sounds, as emotion influences auditory perception.

In an externalization task, twenty-three patients with schizophrenia and twenty-five healthy controls were exposed to neutral and emotional sounds processed to be perceived as: internalized (diotic) or externalized (filtered with either an anechoic head-related transfer function -HRTF- or a binaural room impulse response -BRIR). Participants had to indicate whether the sound source was perceived inside or outside their head. Exploratory analyses also examined the relationships between externalization, reality monitoring, and symptom severity. Compared to controls, patients with schizophrenia rated the filtered sounds (HRTF, BRIR) as less externalized (p_bonf_ < .001) and the diotic sounds as more externalized (p_bonf_ = 0.004), regardless of emotional content. No significant correlation was found between externalization and reality monitoring. In patients, greater symptom severity was associated with reduced externalization of sounds simulated as originating outside the head.

These findings suggest an abnormal perception of sound sources in patients with schizophrenia, who confuse sounds inside and outside the head to a greater extent. Further research is needed to elucidate the relationship between sound externalization and symptoms such as hallucinations.

## 1. Introduction

Schizophrenia, affecting approximately 1% of the population worldwide, is characterized by disruptions in perceptual, cognitive and social abilities that impair daily functioning (Jauhar et al., 2022). A central cognitive disturbance in schizophrenia is the disruption of the self, marked by difficulties in attributing one’s thoughts, actions, emotions, and experiences to oneself, and a tendency to attribute them to others or to external sources. In particular, patients with schizophrenia have difficulties in reality monitoring, the ability to distinguish between their imagination and perceived events (Brunelin et al., 2007; Damiani et al., 2022). This impairment is more severe in patients with auditory hallucinations (Waters et al., 2012), who display higher misattribution of internal (imagined) stimuli as coming from an external source. The cognitive underpinnings of reality monitoring deficit remain incompletely understood, but they are thought to involve deficits in the sense of agency, which determines the subjective origin of productions (self or non-self), and perceptual deficits in spatialization, which determines their subjective source/localization (inner or outer space) (Stephane, 2019). The hypothesis of spatialization deficits is supported by studies reporting bidirectional confusions between internal and external spaces in patients with schizophrenia (Stephane et al., 2010). However, investigations into the distinctions between internal and external spaces have predominantly focused on self-generated productions. Further research is needed to clarify whether the spatialization deficit extends beyond self-generated productions to externally presented stimuli, reflecting broader perceptual deficits in schizophrenia.

Among perceptual deficits in schizophrenia, auditory perception impairments have been consistently reported (Dondé et al., 2017; Fivel et al., 2023), including difficulties in localizing sound sources (Perrin et al., 2010). Research in schizophrenia has primarily focused on the localization of sounds originating from sources located outside the head and received through open ears, in line with most sounds in the natural world. The perception that a sound source is outside the head, referred to as sound externalization (for a review, see Best et al., 2020), is thought to rely on various factors such as reverberation and filtering by the body, particularly the head and pinnae (Catic et al., 2013; Risoud et al., 2018). In experimental settings, it is possible to manipulate sounds delivered through headphones to simulate their perception as being localized inside or outside the head. Indeed, sound sources can be perceived as if originating from inside the head, or internalized, when both the interaural time and level differences are minimal (Brimijoin et al., 2013), for instance when sounds are presented diotically through headphones. In contrast, sound can be perceived as coming from outside the head, or externalized, despite the use of headphones by applying filters such as a head-related transfer function (HRTF) (Wightman and Kistler, 1988) with the addition of simulated reverberation further enhancing the perception of externalization (Leclère et al., 2019). Investigating how individuals with schizophrenia perceive sounds simulated as originating inside or outside the head, by manipulating their degree of externalization, could shed light on spatialization deficits and their role in reality monitoring impairments.

Additionally, the emotional content of auditory information influences both sound localization (Kryklywy et al., 2013) and reality monitoring. While healthy participants are more able to discriminate perceived and imagined words with a negative valence than neutral words (Kensinger and Schacter, 2006), patients with schizophrenia are more likely to misattribute an emotionally negative sentence spoken by themselves as coming from an external source (Pinheiro et al., 2016), suggesting that emotions can lead to source misjudgements. Emotional content may thus influence the degree of perceived externalization.

The present study aimed to investigate how individuals with schizophrenia perceive the localization of sound sources depending on their simulated degree of externalization and emotional content, and how it relates to their reality monitoring abilities and symptoms. We hypothesized that patients with schizophrenia would display greater confusion in sound externalization compared to healthy controls, particularly for sounds conveying negative emotions. Additionally, we explored the relationship between externalization, reality monitoring, and symptom severity. We hypothesized that confusions in externalization would be correlated with misattributions of internal productions as coming from an external source, and with symptom severity. By investigating the externalization of sound sources and its interplay with symptoms, we aimed to advance our understanding of perceptual disturbances in schizophrenia.

## 2. Material and methods

### 2.1. Participants

Twenty-five outpatients diagnosed with schizophrenia according to DSM-5.0 criteria were recruited from Le Vinatier Hospital (Bron, France). Twenty-five healthy controls matched with the patients for age, sex, and education level, were recruited via social networks, universities and hospital staff. Exclusion criteria included medical treatment, current or history of schizophrenia spectrum disorder or other psychiatric conditions and a first-degree relative with schizophrenia or bipolar disorder for healthy controls and, for all participants, history of auditory or neurological impairment, intellectual disability as assessed by Raven’s matrices, and alcohol or substance abuse. Individuals with regular practice of a musical instrument were excluded to prevent potential confounding effects of musical training on sound perception (Micheyl et al., 2006).

The study received approval from a local ethics committee (Comité de Protection des Personnes Mediterranée sud I, France). All participants provided written informed consent. The study was preregistered on February 9, 2021 (NCT04768335; https://clinicaltrials.gov/study/NCT04768335). Recruitment of participants began on March 30, 2021 and ended on December 22, 2022.

### 2.2. Psychopathology assessment

Schizophrenia symptoms were assessed using the Scale for the Assessment of Positive Symptoms (SAPS) and the Positive and Negative Syndrome Scale (PANSS) (Andreasen, 1990; Kay et al., 1988), which assesses the positive, negative, and general psychopathology symptoms.

### 2.3. Procedure

Participants completed perceptual and cognitive tasks on a computer in an isolated room with minimal environmental noise. Sounds were delivered through headphones (HD 250 linear II Sennheiser) at a comfortable level for the participants. Before the experimental trials, participants completed a block of practice trials to ensure they understood the instructions.

### 2.4. Externalization task

The externalization task, programmed using OpenSesame v.3.1., was based on Leclère et al. (2019) and used non-verbal vocalizations from the Montreal Affective Voices battery (Belin et al., 2008). These included vowels /a/ pronounced by male and female actors with neutral or emotional (at the highest intensity level) intonation: anger (growl), fear (scream), disgust (grumble), happiness (laugh), sadness (cry).

The degree of perceived externalization was manipulated in Matlab R2020b by creating three versions of each sound using filters for sound convolution (Leclère et al., 2019): diotic (no filter, assumed to be perceived as originating from the middle the head - internalized), HRTF-filtered (creating an intermediate level of externalization by simulating the effect of specific body characteristics on the sound waves), and filtered by a binaural room impulse response (BRIR; enhancing the degree of perceived externalization by simulating a reverberant environment).

To generate binaural diotic sounds (i.e., stereo sounds), the original monaural vocalizations (sampled at 44100Hz) were duplicated across the two channels (left, right). For binaural filtered sounds, the original sounds were convolved with a HRTF or BRIR measured by Hummersone et al. (2011), using a large cinema-style lecture theatre for the BRIR (room C). To introduce variability in sound source localization in the horizontal plane compared to the diotic sounds (perceived in front/the center), we used HRTFs and BRIRs measured in two left directions relative to the listener (−90°, −60°). Right sources (60°, 90°) were simulated from the left sources by inverting the right and left channels. As azimuths were variability factors, they were not included in the statistical analysis as experimental factors. All sounds were equalized in level by applying the same gain to their left and right channels so that the signal power averaged across the left and right channels was the same for all vocalizations in all conditions. A total of 288 sounds were created and presented, combining 16 sounds * 3 types of sound processing (BRIR, HRTF, diotic) * 6 emotions (neutral, anger, fear, disgust, happiness, sadness).

After each sound, participants were asked to make a binary choice using a keyboard as to the sound source externalization (inside or outside the head). They were given 3 seconds to respond, after which the next sound was presented. Externalization ratings were calculated for each condition as the proportion of trials where the source was perceived outside the head. Participants were expected to have high externalization ratings for BRIR sounds, low ratings for diotic sounds, and intermediate ratings for HRTF sounds (Leclère et al., 2019).

### 2.5. Reality monitoring task

The reality monitoring task, programmed with PsychoPy3 v2020.2.10, was based on the Hear-Imagine task by Brunelin et al. (2008). The task consisted of a presentation phase immediately followed by a test phase. In the presentation phase, 24 neutral French words were visually presented one by one on a computer screen for 3 seconds, preceded by an instruction sentence. Participants were instructed to either listen to the words delivered through headphones or imagine hearing them. The words were pronounced by a neutral male voice. In the test phase, 36 words including the 24 previously presented words and 12 new words, were presented one by one on the computer screen. Participants had to determine whether each word had been presented or not, and if presented, whether it had been heard or imagined. The number of correct source attributions was measured for each source (range 0-12): imagined, heard, and new. Two types of source misattributions were computed: the number of imagined words incorrectly recognized as heard (range 0–12) and the number of heard words incorrectly recognized as imagined (range 0–12).

### 2.6. Analyses

The statistical analyses were conducted using Jamovi (version 2.3.28) and R (version 2023.03), with a significance threshold set at p < 0.05. The sociodemographic data were compared between groups using Fisher’s exact tests for categorical variables and Mann-Whitney U tests for quantitative variables.

As data did not follow a normal distribution, generalized linear models (GLM) with gamma family and identity link function were used. The GLM was used to investigate the effect of group (patients vs. healthy controls), type of sound processing (BRIR, HRTF, diotic), emotional content (neutral, sadness, happiness, anger, fear, disgust), and their interactions on the externalization ratings. Two GLMs were performed on reality monitoring performance to assess whether correct responses were associated with group and source (imagine, hear, new) and whether misattribution errors were associated with group and type of confusion (imagine-to-hear, hear-to-imagine). When a significant association was observed, post-hoc comparisons (contrasts) were conducted with Bonferroni correction.

The relationship between externalization ratings and reality monitoring performance (correct source attributions and source misattributions) was investigated using Spearman correlation. For exploratory analyses, Spearman correlation coefficients were used to assess the relationship with externalization ratings for each type of sound processing and patient’ symptomatology (PANSS negative subscale, PANSS positive subscale, and SAPS hallucination subscale). Bonferroni correction was applied.

## 3. Results

### 3.1. Participant characteristics

The analyses were performed on 47 participants (23 patients with schizophrenia and 24 healthy controls), with data from 3 participants excluded due to missing responses on more than 50% of the trials in the externalization task (reaction times > 3 seconds). The participants’ sociodemographic and clinical characteristics are described in **Table 1**. No significant group differences were found for age, education, or sex.

**Table 1.**
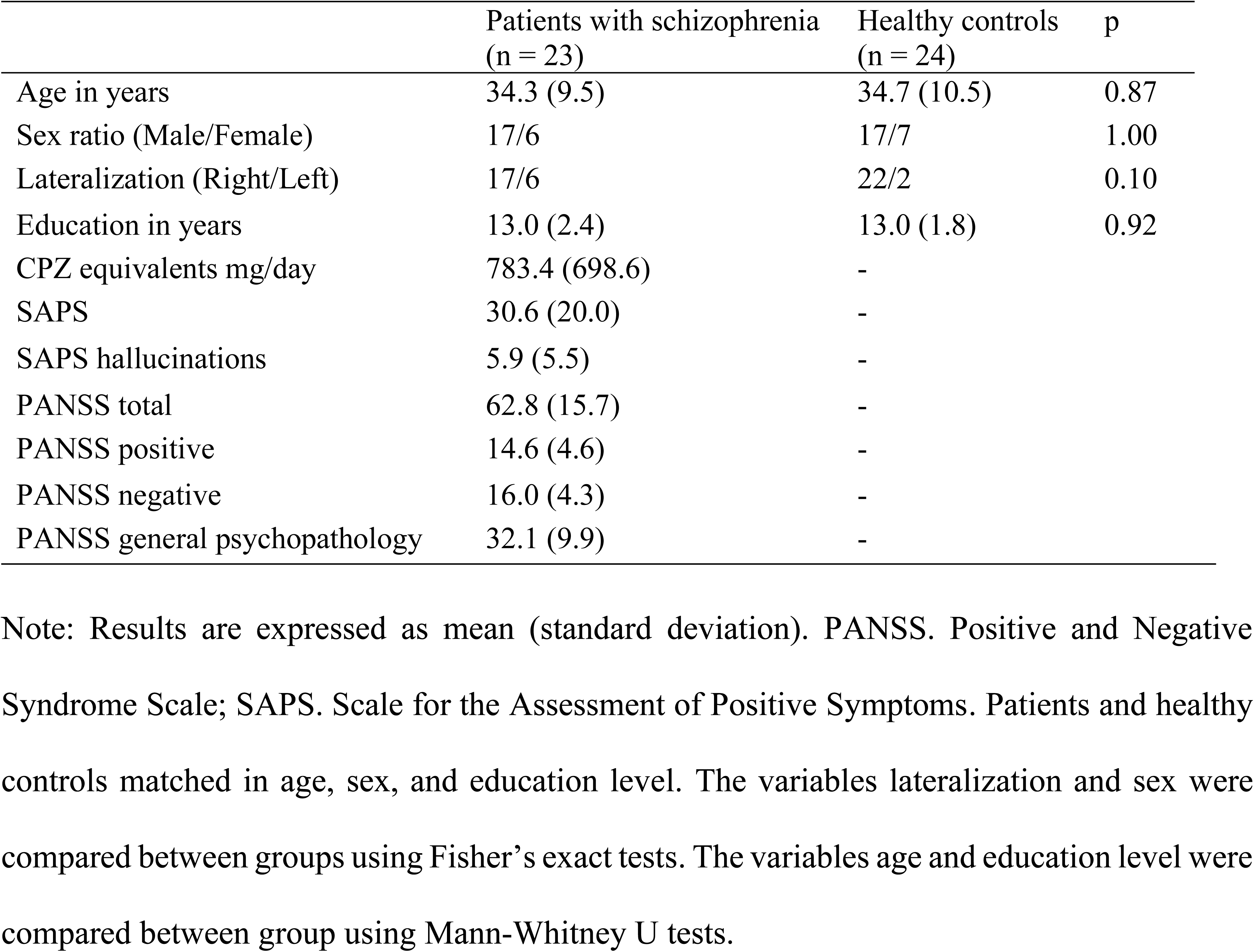
Demographic and clinical characteristics of participants.

### 3.2. Externalization ratings across the 3 types of sound processing

The GLM on externalization ratings revealed a significant group * type of sound processing interaction (β = −29.75, 95% CI [-38.08, −21.46], p < .001, Supplementary Table 1). Patients with schizophrenia rated diotic sounds as significantly more externalized compared to controls (p_bonf_ = 0.004). Conversely, they rated BRIR (p_bonf_ < .001) and HRTF sounds (p_bonf_ < .001) as significantly less externalized compared to controls (**Table 2**, Figure 1). Within each group, there were no significant differences in externalization ratings between BRIR and HRTF sounds (p_bonf_ = 1.00 for both groups).

**Table 2.** Externalization ratings in patients with schizophrenia and healthy controls for each type of sound processing, pooled for emotional content.

|  | Patients with<br>schizophrenia<br>(n = 23) | Healthy controls<br>(n = 24) |  |
| --- | --- | --- | --- |
| BRIR sounds | 70.07 (21.46) | 91.26 (14.73) | *** |
| HRTF sounds | 64.12 (21.13) | 86.40 (21.13) | *** |
| Diotic sounds | 23.36 (20.87) | 14.88 (20.30) | ** |

Note: Results are expressed as mean (standard deviation). HRTF. Head Related Transfer Function; BRIR. Binaural Room Impulse Response. Post hoc of contrasts of the Generalized Linear Model with Bonferroni correction: $**p < .01$ , $***p < .001$ .

**Figure 1.**
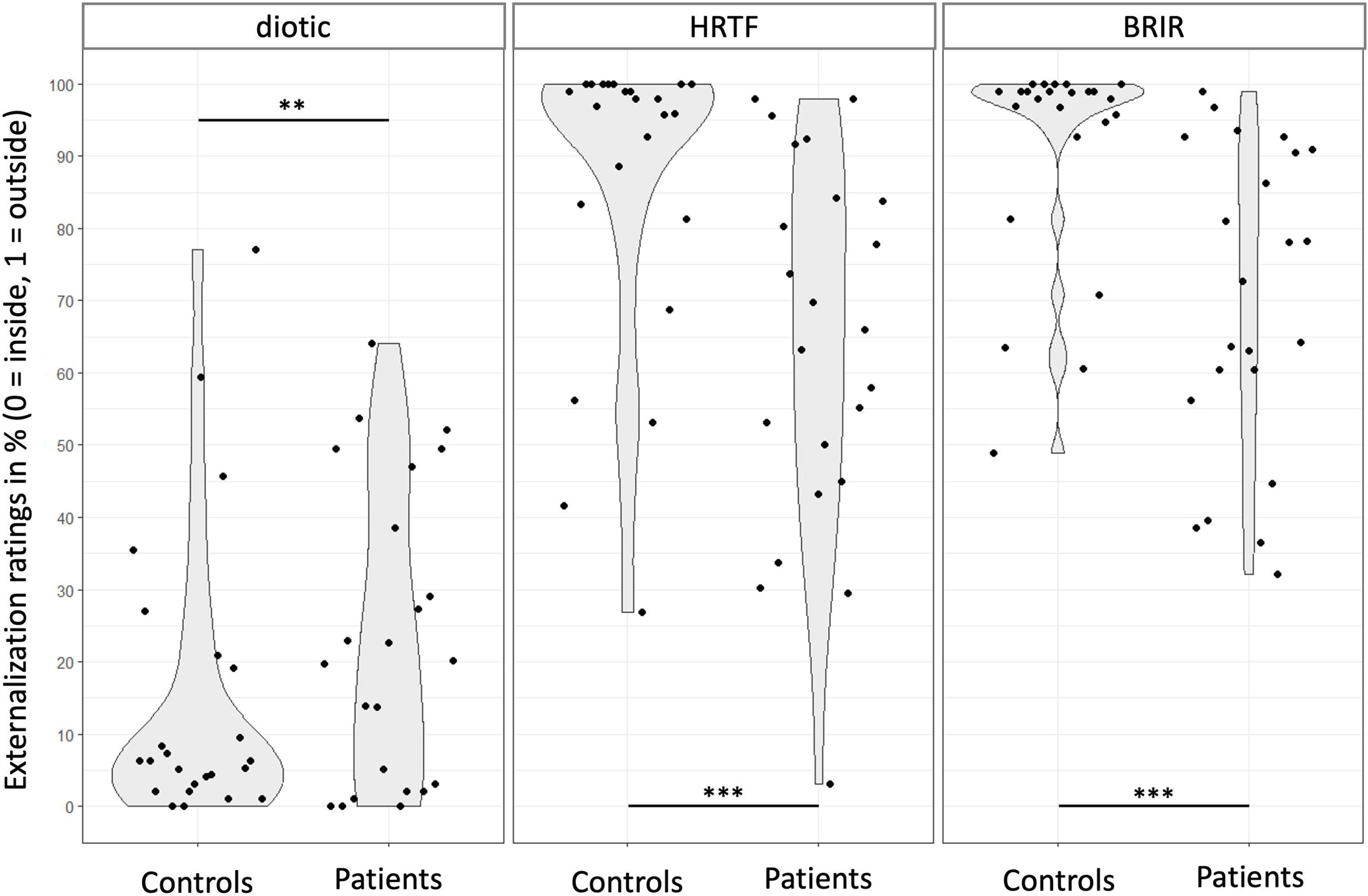
Degree of perceived externalization (sound source perceived as outside the head) for the three types of sound processing (diotic; HRTF: Head Related Transfer Function; BRIR: Binaural Room Impulse Response) in healthy controls (n = 24) and patients with schizophrenia (n = 23). *\*\*\*p <.001; **p <0.01*.

The GLM also revealed significant main effects of group (β = −11.58, 95% CI [-15.15, −8.02], p < .001), type of sound processing (β = 61.50, 95% CI [57.37, 65.68], p < .001) and emotional content (β = 11.86, 95% CI [5.73, 18.01], p < 0.001). Compared to the neutral sounds, sounds conveying fear (p_bonf_ = 0.001) or anger (p_bonf_ = 0.002) were associated with a significantly higher level of perceived externalization, regardless of the type of sound processing or group (**Table 3**, Figure 2). The externalization ratings for each type of sound processing and emotional content in participants are displayed in Supplementary Figure 1.

**Table 3.** Externalization ratings across the emotional content of sounds.

|  | Neutral | Anger | Fear | Disgust | Happiness | Sadness |
| --- | --- | --- | --- | --- | --- | --- |
| Global | 51.78<br>(21.58) | 63.63<br>(17.48) | 63.99<br>(15.86) | 56.60<br>(17.48) | 56.06<br>(18.03) | 58.94<br>(14.98) |
| BRIR | 72.03<br>(31.48) | 86.12<br>(22.13) | 85.52<br>(21.35) | 79.49<br>(24.43) | 78.97<br>(25.31) | 83.20<br>(22.65) |
| HRTF | 68.90<br>(32.79) | 78.74<br>(28.05) | 81.10<br>(24.26) | 72.53<br>(29.13) | 74.43<br>(29.03) | 77.52<br>(26.72) |
| Diotic | 14.40<br>(21.78) | 26.01<br>(27.72) | 25.35<br>(27.38) | 17.77<br>(24.39) | 14.78<br>(19.83) | 16.09<br>(24.13) |

Note: Results are expressed as mean (standard deviation). HRTF. Head Related Transfer Function; BRIR. Binaural Room Impulse Response.

**Figure 2.**
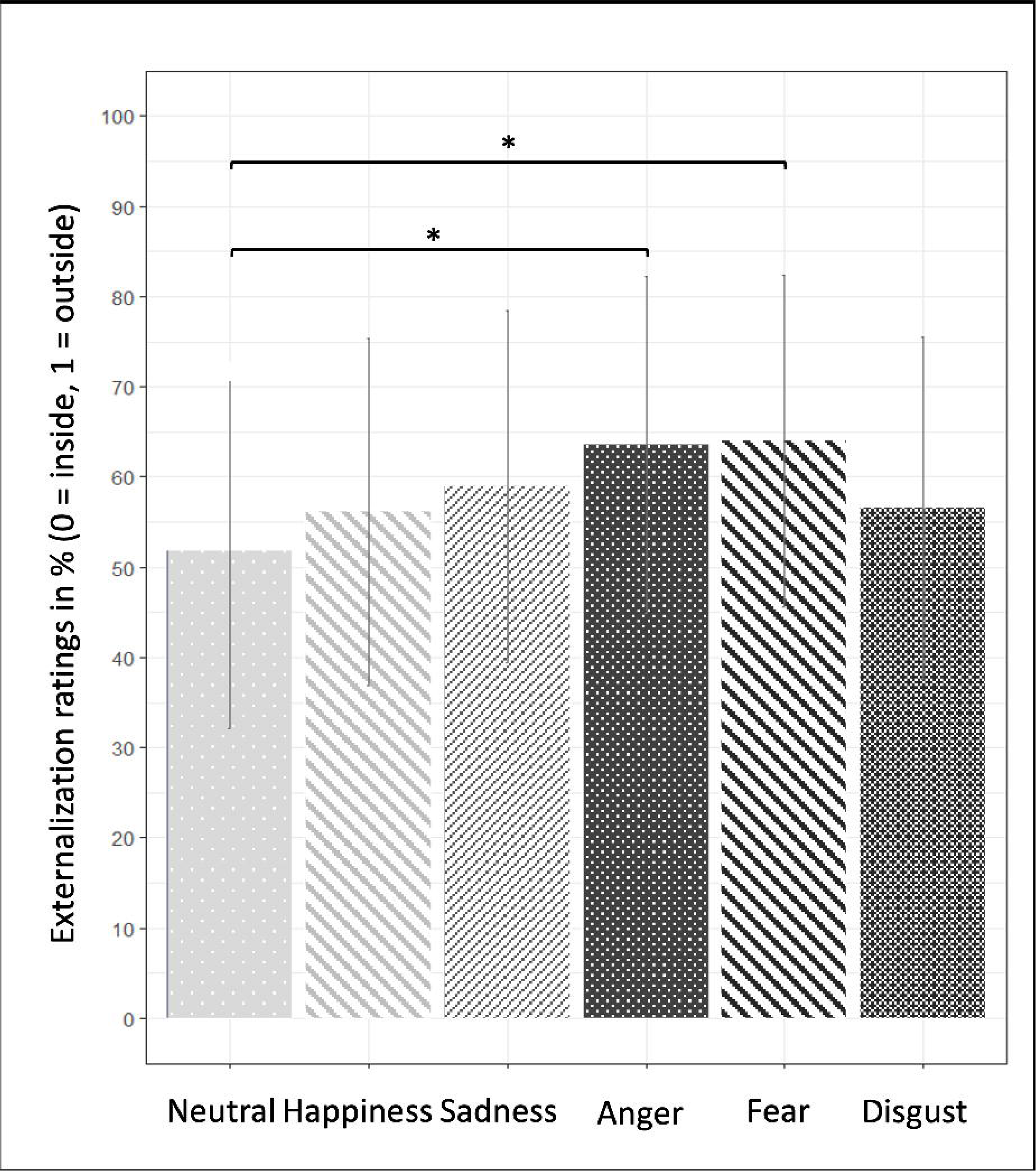
Degree of perceived externalization (sound source perceived as outside the head) across the six emotional contents of sounds, averaged across participant groups (n = 47) and types of sound processing. Results are presented as mean in percent ± one standard deviation. *\*p <.05*.

### 3.3. Reality monitoring task

The GLM on the correct source attributions indicated a significant group * source interaction (β = −2.67, 95% CI [-4.61, −0.73], p = 0.008; Figure 3, Supplementary Table 2). Patients with schizophrenia less accurately recognized the source of imagined words compared with healthy controls (p_bonf_ = 0.04). No significant between-group differences were found for heard or new words (p_bonf_ = 1.00).

**Figure 3.**
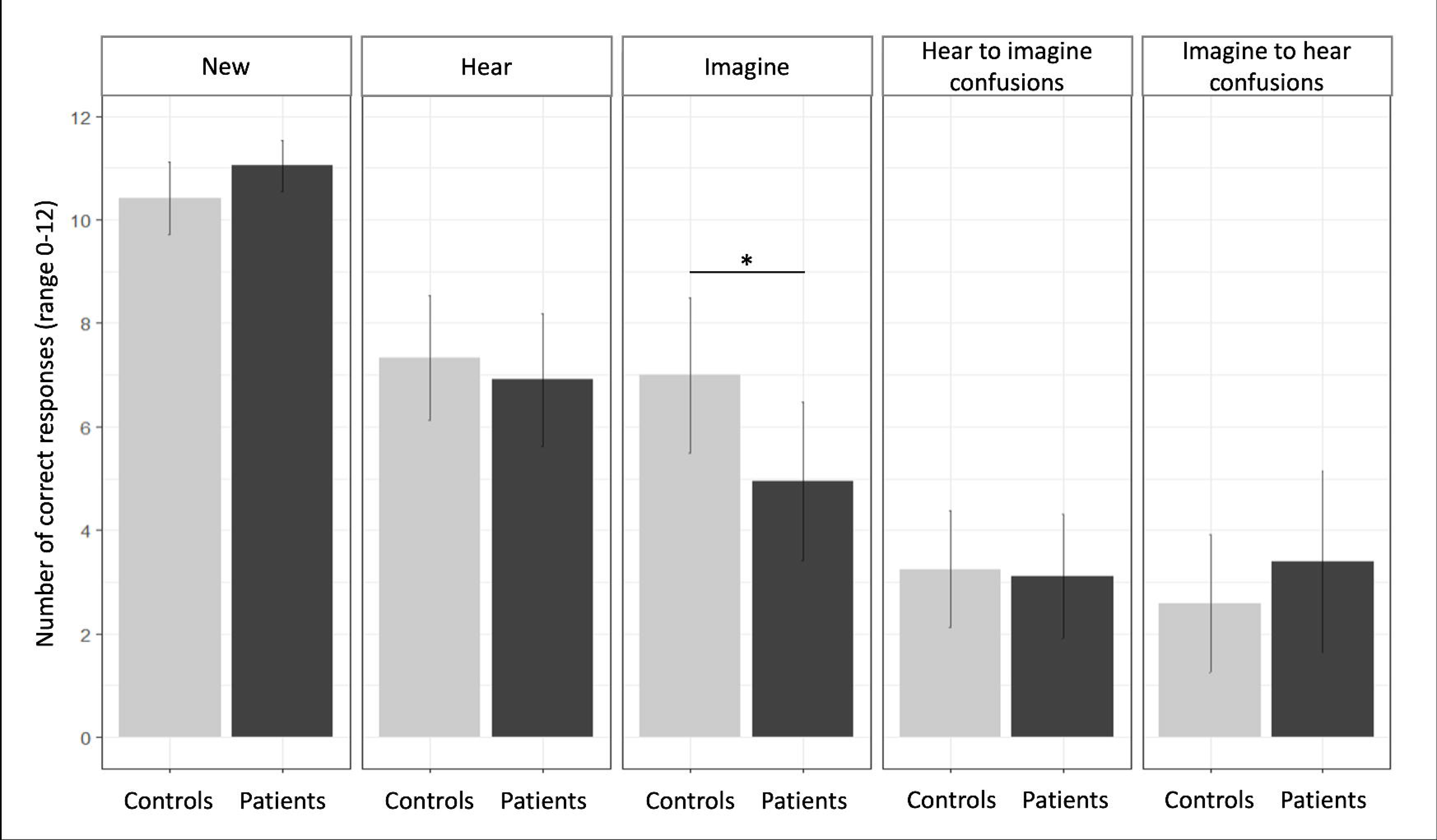
Reality monitoring performance presenting the number of correct source attributions for imagined, heard and new words and the number of source misattributions (heard words recognized as imagined and imagined words recognized as heard) in healthy controls (n = 24) compared with patients with schizophrenia (n = 23). Results are presented as mean ± standard deviation. *\*p <.05*.

The GLM on the source misattributions revealed no significant interaction between group * type of source misattribution (Supplementary Table 3).

### 3.4. Relationship between externalization and reality monitoring

No correlation was found between externalization ratings and reality monitoring performance, neither within healthy controls and patients separately, nor across the two groups.

### 3.5. Relationship between externalization and symptomatology

A significant negative correlation was found between scores on the PANSS negative subscale and externalization ratings of BRIR sounds (r = −0.47, p = 0.02), whereas no significant correlations were found for diotic and HRTF sounds. In exploratory analyses, the relationships between the externalization of emotional sounds and symptoms were further examined (Supplementary Table 4 and Supplementary Figure 2). The PANSS negative subscale was negatively correlated with the externalization ratings of BRIR (r = −0.44, p = 0.03) and HRTF (r = −0.43, p = 0.03) neutral sounds, and BRIR sounds conveying disgust (r = −0.49, p = 0.01). PANSS positive subscale was correlated positively with the externalization ratings of neutral (r = 0.49, p = 0.03) and happiness diotic sounds (r = 0.49, p = 0.02), and negatively with the externalization ratings of BRIR sounds conveying anger (r = −0.42, p = 0.04). Moreover, SAPS hallucination scores were negatively correlated with the externalization ratings of BRIR (r = - 0.42, p = 0.04) and HRTF (r = −0.43, p = 0.03) sounds conveying fear. None of these correlations reach statistical significance after Bonferroni correction (threshold at α=0.05/18 = 0.002).

## 4. Discussion

For the first time, we investigated the externalization of sound sources using neutral and emotional vocalizations in patients with schizophrenia and healthy controls. Our results highlighted significant differences in sound externalization between the two groups, but no correlation was found with reality monitoring abilities. Sounds conveying emotions of fear or anger were perceived as more externalized in both groups. Exploratory analyses suggested potential correlations between externalization and patients’ symptomatology, but none reached statistical significance after Bonferroni correction.

### 4.1. Emotional content of sound modulates the perception of externalization

To the best of our knowledge, research on the impact of emotion on sound localization is limited, with most studies focusing on reaction time rather than performance. The current study brings new data in the research field of externalization by suggesting that emotional valence modulates the degree of perceived externalization. Sounds conveying negative emotions, such as anger and fear, were rated as more externalized than neutral sounds, regardless of the group and the type of sound processing. These results may reflect a tendency to attribute danger to the external environment. Fearful or angry voices may signal threat and trigger a response to danger, leading to a bias in processing emotional stimuli, thereby predicting an external cause for these emotions.

Interestingly, previous studies revealed that auditory information conveying negative emotional valence reduces self-agency compared to neutral or positive information (Christensen et al., 2019; Yoshie and Haggard, 2013). Participants are less likely to attribute action or sound manifestations to themselves when they convey fear or anger. Similarly, healthy controls make more external attributions to negative than to positive events (Lyon et al., 1999). In relation to our results, higher externalization of sources conveying fear or anger could serve as a proxy for self-agency disturbances in a lower-level processing.

### 4.2. Externalization of sound sources in schizophrenia

Healthy controls showed the expected pattern of externalization, with BRIR and HRTF sounds leading to sources perceived as more externalized, and diotic sounds leading to more internalized sources. In contrast, patients with schizophrenia showed lower externalization for BRIR and HRTF sounds compared to controls, and higher externalization for diotic sounds. These confusions could reflect a general deficit of sound localization, as externalization and localization processes share common underlying neurological bases (Callan et al., 2013; Hunter et al., 2003). The bidirectional confusions observed in patients may arise from abnormal processing of binaural cues. Binaural cues play a critical role in externalization perception, with less temporal variations in interaural differences leading to reduced perceived externalization, the temporal variations being associated with the effect of reverberation (Catic et al., 2015). Patients with schizophrenia have difficulty in using these cues to localize sounds in space (Matthews et al., 2013), which may explain the observed confusions – namely, the slightly more externalized perception of diotic sounds and the more internalized perception of dichotic reverberant sounds. However, achieving full externalization might require more than binaural cues; it also involves the processing of body filtering (simulated by HRTF) and room reverberation (simulated by BRIR) (Boyd et al., 2012), even if we cannot exclude the possibility that they influence externalization only by causing the temporal variations in binaural cues. Further investigating these acoustical cues in schizophrenia may help to understand which individual factors or combinations contribute to these perceptual confusions and to some extent to associated cognitive processes such as spatial self-representation, since externalization appears indispensable to the ability to situate elements of one’s environment in relation to oneself (Heine et al., 2021).

### 4.3. Externalization of sound sources in relation to the severity of symptoms

The wide variability in externalization ratings in patients with schizophrenia leads us to hypothesize that symptom severity could account for this variability. Higher levels of negative symptoms were associated with lower externalization of neutral sources simulated outside the head (HRTF and BRIR sounds). This finding is consistent with a model suggesting that auditory processing deficits in schizophrenia contribute to poor social and cognitive functioning via negative symptom severity (Thomas et al., 2017). We could speculate that difficulty in perceiving external sources outside the head may contribute to social functioning deficits commonly seen in schizophrenia, such as weak bodily self-representation (Gallese and Ferri, 2014) and impaired social cognition (Lin et al., 2013). However, the absence of a specific scale evaluating negative symptoms prevent us from investigating which subtype of negative symptoms would be most strongly related to externalization confusions. Moreover, we found the hint of an association between hallucination severity and lower externalization of fearful sounds simulated outside the head. Brain activity of patients with auditory hallucinations respond differently to emotional words compared to patients without hallucinations (Escartí et al., 2010) and healthy controls (Sanjuan et al., 2007), especially to negative words. Notably, abnormal increased amygdala activation in resting state and in response to emotional stimuli has been observed in patients with hallucinations. Therefore, auditory perception of fear may contribute to externalization confusions especially in patients with hallucinations. Nevertheless, these hypotheses remain highly speculative, as the observed correlations did not survive correction for multiple comparisons.

### 4.4. Relationship between externalization of sound sources and reality monitoring

Our exploratory investigation found no evidence of a relationship between externalization and reality monitoring. Stephane (2019) suggested that reality monitoring involves two independent processes: self-agency and spatialization. The absence of a link in our study may indicate a more critical role for self-agency than spatialization in reality monitoring. This is consistent with a recent study reporting a significant relationship between self-agency and reality monitoring abilities, although spatialization was not assessed (Subramaniam et al., 2018). Our findings might also be explained by the fact that we did not find evidence of biases in patients’ reality monitoring performance (i.e., patients did not exhibit more ‘imagine-to-hear’ confusions than controls, which is typically observed in schizophrenia (Brookwell et al., 2013; Brunelin et al., 2006)) although we found that imagined words are less recognized in patients compared to healthy controls. This may be explained by our sample including patients with less severe symptoms than those in previous studies (total PANSS score of 62.8 versus 80.9 in Dondé et al., 2019a). In addition, half of our patient sample (13/23) had no hallucinations, and the presence of hallucinations has been associated with greater reality monitoring confusions (Brunelin et al., 2006; Woodward et al., 2007). Nevertheless, our findings do not allow us to conclude to an absence of relationship between externalization and reality monitoring abilities. Further research is needed to explore the potential implication of externalization in reality monitoring, particularly in patients with auditory hallucinations.

### 4.5. Limitations and Perspectives

Our study presents several limitations that should be acknowledged. First, our results contradict previous studies suggesting that reverberation added in BRIR sounds enhances externalization compared to HRTF (Leclère et al., 2019). The absence of evidence for this effect may indicate that HRTF alone were sufficient to create an external source experience in healthy controls (with > 84% of HRTF sounds perceived outside the head). Another explanation is the use of a binary forced-choice response (outside or inside the head) to evaluate externalization. While this approach, used in prior research (Brimijoin et al., 2013; Ohl et al., 2010), may better correspond to the percept being measured, it may make it more challenging to distinguish true from guessed responses, despite explicit instructions emphasizing that there were no right or wrong answers, only individual perception. Some other studies have used a continuous scale (Kates et al., 2018; Leclère et al., 2019), which allows for greater response variability but may introduce a reference to distance, potentially misleading to measure externalization (Best et al., 2020). Second, we did not control for attentional engagement, as reduced attention may impair performance, particularly in patients with schizophrenia (Carter et al., 2010). However, we attempted to mitigate this by using a 3 second cut-off for response times, as such delays may indicate attentional lapses. Third, the small sample size limits statistical power, particularly for exploring the effects of emotion on externalization or the relationship between externalization and reality monitoring. Additionally, the absence of a comparative group of patients precludes conclusions about whether externalization confusions are specific to schizophrenia. Finally, our study was not designed to investigate differences between patients with and without hallucinations, or the association with specific types of hallucinations. Auditory hallucinations in schizophrenia can be experienced inside or outside the head (Copolov et al., 2004), with distinct neural correlates for each (Looijestijn et al., 2013). Further studies should explore whether differences in externalization are linked to the nature of hallucinations.

Despite these limitations, our findings open new perspectives for therapeutic interventions in schizophrenia. Auditory sensory training has shown benefits in improving auditory perception, cognitive functioning, and reducing symptoms in patients (Dondé et al., 2019b). Given that sound externalization can be improved through training with HRTF- or BRIR-filtered sounds (Mitruț et al., 2015; Klein et al., 2017), future research should investigate whether such training could address externalization deficits in schizophrenia, potentially improving symptoms.

### 4.6. Conclusions

This study highlights abnormal perception of sound sources in patients with schizophrenia, with increased internal/external confusions. It also provides new insights into externalization, highlighting the role of negative emotions such as fear and anger in increasing perceived externalization. Future research should further investigate sound externalization in schizophrenia and its relation to patients’ symptomatology and higher cognitive processes such as reality monitoring.

## Data Availability

All data produced in the present study are available upon reasonable request to the authors

## Acknowledgement

The authors thank Gabrielle CHESNOY for her help with the study design, participant recruitment and patient inclusion. We thank Asako TAKAMATSU for her help with the participant recruitment and data collection. We also thank all the participants of the study.

## Fundings

This research is financially supported by the Scientific Council of CH Le Vinatier (#CSRM01).

## Disclosure Statement

The authors declare no conflict of interest.

## Author Contributions

FH: conceptualization; help in the formal analysis; supervision; validation; writing -original draft preparation; writing – review and editing. MM: conceptualization; help in the formal analysis; supervision; validation; writing -original draft; writing – review and editing. FP: conceptualization; help in the formal analysis; methodology; validation. NG: conceptualization; help in the formal analysis; methodology; validation. ML: conceptualization; help in the formal analysis; methodology; validation; writing – review and editing. LF: data curation; writing – original draft preparation; writing – review and editing.

## Notes

### Competing Interest Statement

The authors have declared no competing interest.

### Clinical Trial

NCT04768335

### Author Declarations

Ethics committee of Mediterranee sud I (France) gave ethical approval for this work.

