## Supplementary Table 1 for "Is it inside my head? Characterization of sound externalization in schizophrenia"

**Supplementary Material for the paper “Is it inside my head? Characterization of sound externalization in schizophrenia”**

Laure FIVEL, Mathieu LAVANDIER, Nicolas GRIMAULT, Fabien PERRIN*, Marine MONDINO*, Frédéric HAESEBAERT*

* *co-last authorship*

**Supplementary Table 1. Results of the Generalized Linear Model (GLM) analyzing Externalization ratings by Group, Sound Type, and Emotion**

| **Model** | **df** | **AIC** | **LogLik** | | **χ^2^** | **p** |
| --- | --- | --- | --- | --- | --- | --- |
| Group * Type of sound processing * Emotion type | 10 | 7910.82 | -3918.41 | | 0.57 | 1.000 |
| **Estimated effects** |  |  | **95% confidence interval** | |  |  |
|  | **Estimate** | **SE** | **Lower** | **Upper** | **z** | **p** |
| **(Intercept)** | **158.3786** | **0.909** | **156.61** | **160.17** | **174.15932** | **< .001** |
| **patients - controls** | **-11.5845** | **1.819** | **-15.15** | **-8.02** | **-6.36939** | **< .001** |
| **BRIR - diotic** | **61.5047** | **2.120** | **57.37** | **65.68** | **29.00839** | **< .001** |
| **HRTF - diotic** | **56.1398** | **2.077** | **52.09** | **60.23** | **27.03186** | **< .001** |
| **anger - neutral** | **11.8629** | **3.131** | **5.73** | **18.01** | **3.78880** | **< .001** |
| disgust - neutral | 4.8599 | 3.063 | -1.14 | 10.87 | 1.58644 | 0.113 |
| **fear - neutral** | **12.2609** | **3.136** | **6.12** | **18.42** | **3.90984** | **< .001** |
| happiness - neutral | 4.2470 | 3.061 | -1.75 | 10.25 | 1.38761 | 0.166 |
| **sadness - neutral** | **7.1833** | **3.091** | **1.13** | **13.25** | **2.32373** | **0.020** |
| **patients - controls ✻ BRIR - diotic** | **-29.7599** | **4.240** | **-38.08** | **-21.46** | **-7.01805** | **< .001** |
| **patients - controls ✻ HRTF - diotic** | **-30.7400** | **4.154** | **-38.89** | **-22.61** | **-7.40080** | **< .001** |
| patients - controls ✻ anger - neutral | 1.2204 | 6.262 | -11.06 | 13.50 | 0.19488 | 0.846 |
| patients - controls ✻ disgust - neutral | 3.7192 | 6.127 | -8.29 | 15.74 | 0.60703 | 0.544 |
| patients - controls ✻ fear - neutral | 4.4354 | 6.272 | -7.86 | 16.74 | 0.70719 | 0.480 |
| patients - controls ✻ happiness - neutral | -3.4326 | 6.121 | -15.44 | 8.57 | -0.56076 | 0.575 |
| patients - controls ✻ sadness - neutral | 1.8869 | 6.183 | -10.24 | 14.01 | 0.30520 | 0.760 |
| BRIR - diotic ✻ anger - neutral | 2.4822 | 7.317 | -11.86 | 16.85 | 0.33922 | 0.735 |
| HRTF - diotic ✻ anger - neutral | -1.7434 | 7.168 | -15.80 | 12.32 | -0.24321 | 0.808 |
| BRIR - diotic ✻ disgust - neutral | 4.0658 | 7.143 | -9.94 | 18.09 | 0.56919 | 0.569 |
| HRTF - diotic ✻ disgust - neutral | 0.2308 | 7.001 | -13.50 | 13.97 | 0.03296 | 0.974 |
| BRIR - diotic ✻ fear - neutral | 2.5371 | 7.303 | -11.78 | 16.87 | 0.34740 | 0.728 |
| HRTF - diotic ✻ fear - neutral | 1.2703 | 7.197 | -12.84 | 15.40 | 0.17651 | 0.860 |
| BRIR - diotic ✻ happiness - neutral | 6.5985 | 7.104 | -7.33 | 20.54 | 0.92879 | 0.353 |
| HRTF - diotic ✻ happiness - neutral | 5.1496 | 6.997 | -8.57 | 18.88 | 0.73597 | 0.462 |
| BRIR - diotic ✻ sadness - neutral | 9.4704 | 7.180 | -4.60 | 23.57 | 1.31891 | 0.188 |
| HRTF - diotic ✻ sadness - neutral | 6.8866 | 7.055 | -6.94 | 20.74 | 0.97607 | 0.329 |
| patients - controls ✻ BRIR - diotic ✻ anger - neutral | 1.0595 | 14.635 | -27.65 | 29.76 | 0.07240 | 0.942 |
| patients - controls ✻ HRTF - diotic ✻ anger - neutral | 3.3188 | 14.336 | -24.80 | 31.44 | 0.23149 | 0.817 |
| patients - controls ✻ BRIR - diotic ✻ disgust - neutral | -1.3636 | 14.286 | -29.39 | 26.66 | -0.09545 | 0.924 |
| patients - controls ✻ HRTF - diotic ✻ disgust - neutral | -2.3163 | 14.001 | -29.78 | 25.15 | -0.16543 | 0.869 |
| patients - controls ✻ BRIR - diotic ✻ fear - neutral | 0.9609 | 14.606 | -27.69 | 29.61 | 0.06579 | 0.948 |
| patients - controls ✻ HRTF - diotic ✻ fear - neutral | 2.5406 | 14.393 | -25.69 | 30.78 | 0.17651 | 0.860 |
| patients - controls ✻ BRIR - diotic ✻ happiness - neutral | 4.9518 | 14.209 | -22.92 | 32.82 | 0.34850 | 0.728 |
| patients - controls ✻ HRTF - diotic ✻ happiness - neutral | -0.1522 | 13.994 | -27.61 | 27.29 | -0.01088 | 0.991 |
| patients - controls ✻ BRIR - diotic ✻ sadness - neutral | 0.0705 | 14.361 | -28.10 | 28.24 | 0.00491 | 0.996 |
| patients - controls ✻ HRTF - diotic ✻ sadness - neutral | -3.7539 | 14.111 | -31.44 | 23.92 | -0.26603 | 0.790 |

Note: The table presents the results of the Generalized Linear Model (GLM) with externalization ratings as the dependent variable. A constant of 100 was added to the ratings to allow for analysis with the Gamma family, as non-positive values are not permitted. This adjustment was necessary because some participants recorded zero values when sounds were not perceived outside the head. The upper part of the table presents the Loglikelihood ratio tests with the degrees of freedom (df), Akaike Information Criterion (AIC), LogLikelihood ratio (LogLik), Chi square (χ^2^) and the p-value of the interactions. The lower part of the table presents a summary of the estimated effects, standard errors (SE), confidence intervals (95%), and corresponding statistical metrics. The reference levels are as follows: *healthy* *controls* for the group factor, *diotic sounds* for the sound type factor, and *neutral* for the emotion factor. Statistical significance is set at p-values < .05.
