## Supplementary Table 3 for "Is it inside my head? Characterization of sound externalization in schizophrenia"

**Supplementary Material for the paper “Is it inside my head? Characterization of sound externalization in schizophrenia”**

Laure FIVEL, Mathieu LAVANDIER, Nicolas GRIMAULT, Fabien PERRIN*, Marine MONDINO*, Frédéric HAESEBAERT*

* *co-last authorship*

**Supplementary Table 3. Results of the Generalized Linear Model (GLM) analyzing Reality monitoring performance by Group and Type of Source Misattributions.**

| **Model** | **df** | **AIC** | **LogLik** | | **χ^2^** | **p** |
| --- | --- | --- | --- | --- | --- | --- |
| Group * Source misattribution type | 1 | 395.77 | -192.31 | | 0.19 | 0.660 |
| **Estimated effects** |  |  | **95% Confidence interval** | |  |  |
|  | **Estimate** | **SE** | **Lower** | **Upper** | **z** | **p** |
| **(intercept)** | **102.201** | **0.199** | **101.811** | **102.59** | **514.693** | **<.001** |
| patients - controls | -0.140 | 0.397 | -0.918 | 0.638 | -0.354 | 0.725 |
| Hear to imagine confusions – Imagine to hear confusions | 0.784 | 0.397 | 0.005 | 1.562 | 1.973 | 0.052 |
| patients - controls * Hear to imagine confusions - Imagine to hear confusions | -0.350 | 0.794 | -1.906 | 1.207 | -0.440 | 0.661 |

Note: The table presents the results of the Generalized Linear Model (GLM) with performance as the dependent variable. A constant of 100 was added to the performance scores to allow for analysis with the Gamma family, as non-positive values are not permitted. This adjustment was necessary because some participants recorded zero values when no misattribution was performed. The upper part of the table presents the Loglikelihood ratio tests with the degrees of freedom (df), Akaike Information Criterion (AIC), LogLikelihood ratio (LogLik), Chi square (χ^2^) and the p-value of the interactions. The lower part of the table presents a summary of the estimated effects, standard errors (SE), confidence intervals (95%), and corresponding statistical metrics. The reference levels are as follows: *healthy controls* for the group factor, *imagine to hear misattributions* for the type of source misattributions. Statistical significance is set at p-values < .05.
