## Supplementary Table 4 for "Is it inside my head? Characterization of sound externalization in schizophrenia"

**Supplementary Material for the paper “Is it inside my head? Characterization of sound externalization in schizophrenia”**

Laure FIVEL, Mathieu LAVANDIER, Nicolas GRIMAULT, Fabien PERRIN*, Marine MONDINO*, Frédéric HAESEBAERT*

* *co-last authorship*

**Supplementary Table 4.** Correlations of the symptomatology scores with the externalization ratings.

|  | **Neutral** | | | **Anger** | | | **Fear** | | |
| --- | --- | --- | --- | --- | --- | --- | --- | --- | --- |
|  | Diotic  (n = 48) | HRTF  (n = 48) | BRIR  (n = 48) | Diotic  (n = 48) | HRTF  (n = 48) | BRIR  (n = 48) | Diotic  (n = 48) | HRTF  (n = 48) | BRIR  (n = 48) |
| PANSS negative subscore | r =  -0.074 | r =  -0.439 | r =  -0.445^#^ | r =  0.094 | r =  -0.061 | r =  -0.250 | r =  -0.262 | r =  -0.112 | r =  -0.224 |
| PANSS positive subscore | r =  0.449 | r =  -0.065 | r =  -0.078 | r =  0.119 | r =  -0.318 | r =  -0.420 | r =  0.175 | r =  -0.327 | r =  -0.413 |
| Hallucination (SAPS) | r =  0.206 | r =  -0.243 | r =  -0.101 | r =  0.152 | r =  -0.413 | r =  -0.387 | r =  0.092 | r =  -0.435 | r =  -0.428 |
|  | **Disgust** | | | **Happiness** | | | **Sadness** | | |
|  | Diotic  (n = 48) | HRTF  (n = 48) | BRIR  (n = 48) | Diotic  (n = 48) | HRTF  (n = 48) | BRIR  (n = 48) | Diotic  (n = 48) | HRTF  (n = 48) | BRIR  (n = 48) |
| Negative PANSS subscore | r =  -0.033 | r =  -0.270 | r =  -0.499 | r =  0.884 | r =  0.358 | r =  0.181 | r =  0.078 | r =  -0.389 | r =  -0.402 |
| Positive PANSS subscore | r =  -0.177 | r =  0.331 | r =  -0.140 | r =  0.488 | r =  0.005 | r =  0.029 | r =  0.359 | r =  -0.221 | r =  -0.348 |
| Hallucination (SAPS) | r =  -0.027 | r =  -0.339 | r =  -0.212 | r =  0.089 | r =  -0.361 | r =  -0.161 | r =  0.035 | r =  -0.306 | r =  -0.263 |

Correlations of the patients’ symptomatology scores with the externalization ratings across the three types of processing (Diotic; HRTF: *Head Related Transfer Function*; BRIR: *Binaural Room Impulse Response*) and the emotions. Correlations were assessed with a Spearman test. None of the correlations was significant after Bonferroni correction (α=0.05/18=0.002). PANSS: *Positive and negative syndrome scale*; SAPS: *Scale for the Assessment of Positive Symptoms*.
