## Supplementary Figure 1 for "Is it inside my head? Characterization of sound externalization in schizophrenia"

**Supplementary Material for the paper “Is it inside my head? Characterization of sound externalization in schizophrenia”**

Laure FIVEL, Mathieu LAVANDIER, Nicolas GRIMAULT, Fabien PERRIN*, Marine MONDINO*, Frédéric HAESEBAERT*

* *co-last authorship*

**Supplementary Figure 1.** Externalization ratings (sound source perceived as outside (1) or inside (0) the head) for the three types of sound processing (Diotic; HRTF: Head Related Transfer Function; BRIR: Binaural Room Impulse Response) and the six emotional contents in healthy controls (n = 24, in blue-green) and patients with schizophrenia (n = 23, in pink). Results are displayed as mean in percent ± one standard deviation.

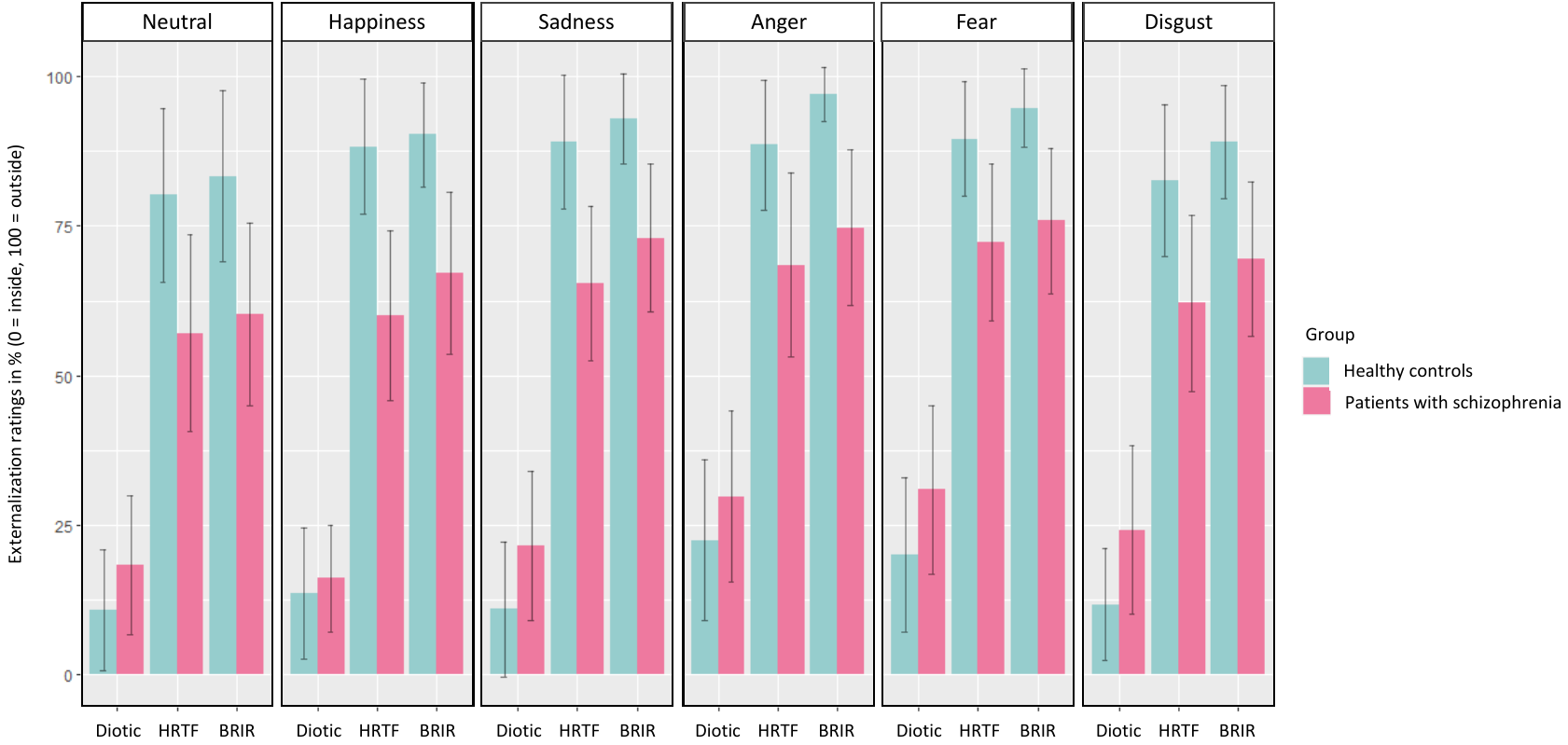
