## Supplementary Figure 2 for "Is it inside my head? Characterization of sound externalization in schizophrenia"

**Supplementary Material for the paper “Is it inside my head? Characterization of sound externalization in schizophrenia”**

**Supplementary Figure 2.** Scatterplots of the patients’ symptomatology scores as a function of the externalization ratings across the three types of processing (Diotic; HRTF: *Head Related Transfer Function*; BRIR: *Binaural Room Impulse Response*). Scatterplots are plotted for each of the correlations leading to a Spearman coefficient with an absolute value above 0.4. Note that none of these correlations was significant after Bonferroni correction (α=0.002). PANSS: *Positive and negative syndrome scale*; SAPS: *Scale for the Assessment of Positive Symptoms*.

**Figure 2A. Scatterplot of the negative symptoms as a function of the externalization ratings**

| Negative PANSS subscore | 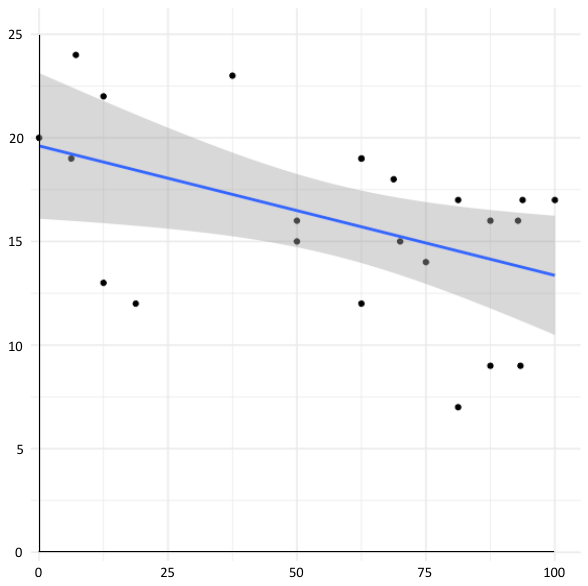  r = -0.439  p = 0.036 | 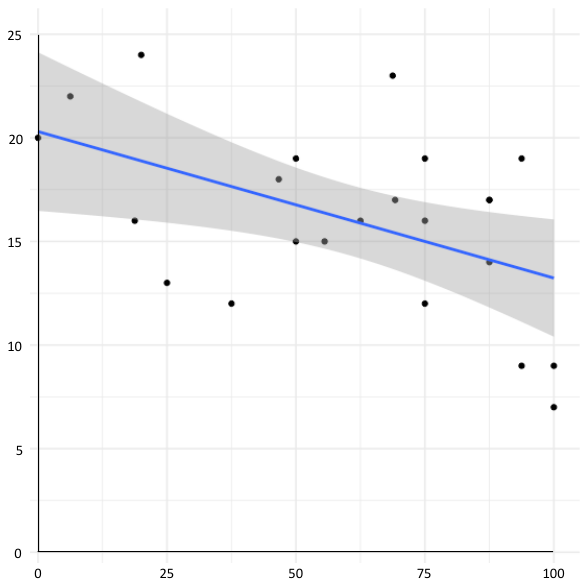  r = -0.445  p = 0.033 | 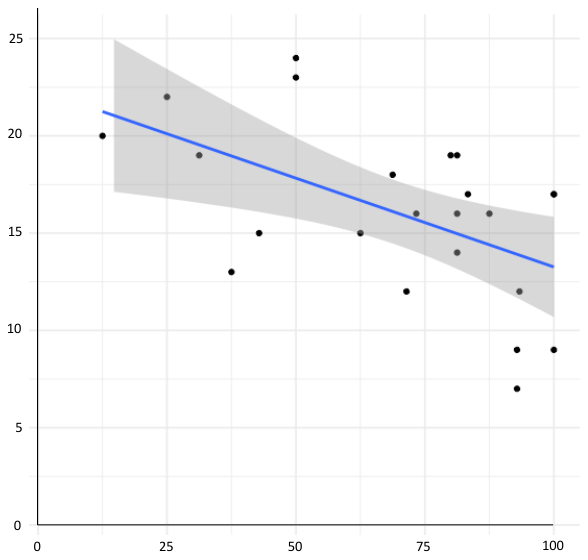  r = -0.499  p = 0.015 |
| --- | --- | --- | --- |
|  | Externalization ratings of neutral HRTF sounds | Externalization ratings of neutral BRIR sounds | Externalization ratings of disgust  BRIR sounds |

**Figure 2B. Scatterplot of the positive symptoms as a function of the externalization ratings**

| Positive PANSS subscore | 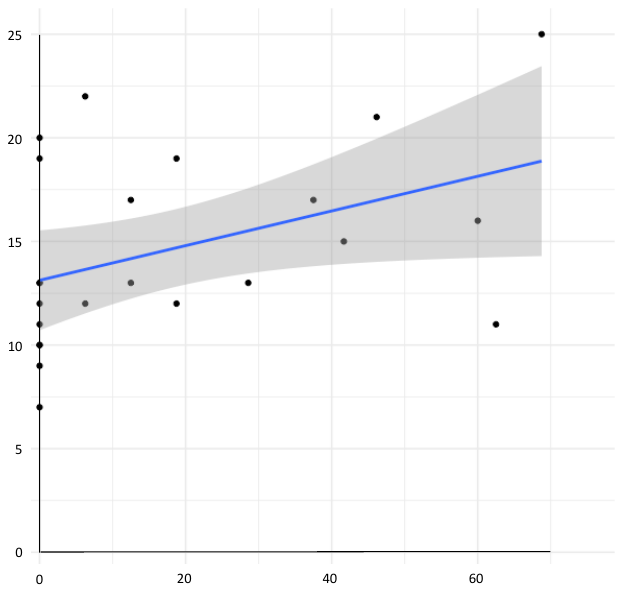  r = 0.449  p = 0.032 | 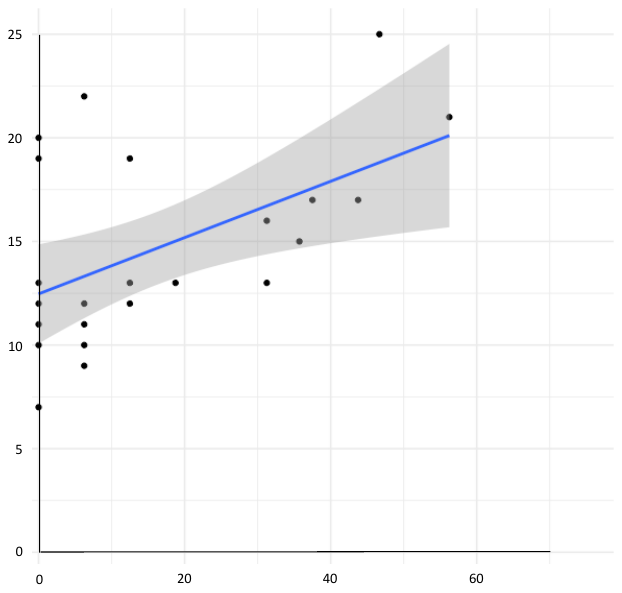  r = 0.488  p = 0.018 | 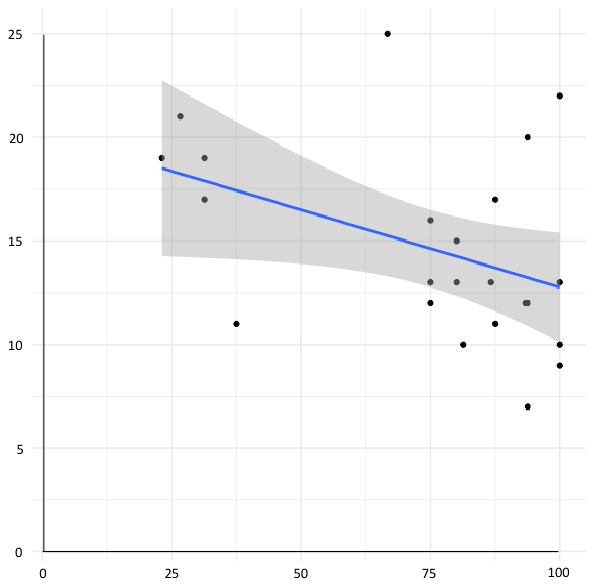  r = -0.420  p = 0.046 |
| --- | --- | --- | --- |
|  | Externalization ratings of neutral  diotic sounds | Externalization ratings of happiness diotic sounds | Externalization ratings of anger  BRIR sounds |

**Figure 2C. Scatterplot of the hallucinations as a function of the externalization ratings**

| SAPS Hallucination subscore | 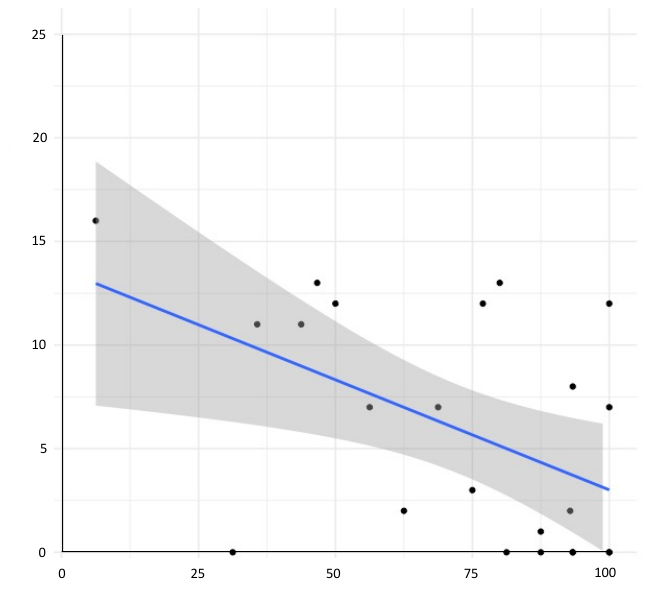  r = -0.435  p = 0.038 | 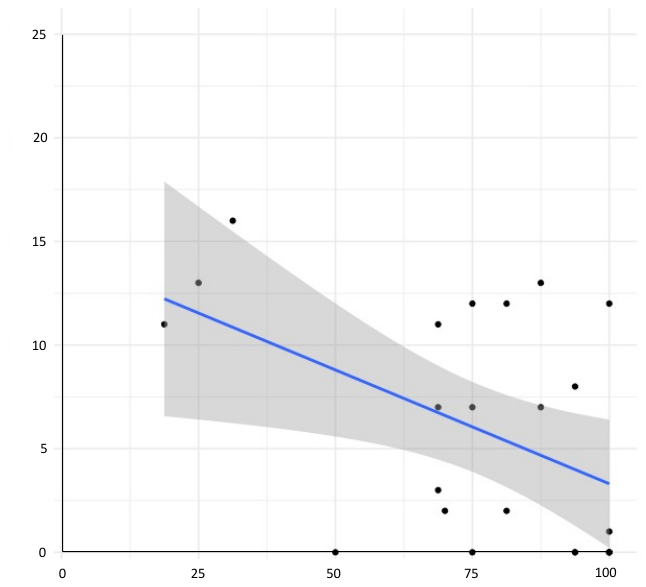  r = -0.428  p = 0.042 |
| --- | --- | --- |
|  | Externalization ratings of fear  HRTF sounds | Externalization ratings of fear  BRIR sounds |
